# Post-GWAS multiomic functional investigation of the *TNIP1* locus in Alzheimer’s disease implicates mediation through *GPX3*

**DOI:** 10.1101/2022.11.04.22277162

**Authors:** Daniel J. Panyard, Lianne M. Reus, Muhammad Ali, Jihua Liu, Yuetiva K. Deming, Qiongshi Lu, Gwendlyn Kollmorgen, Ivonne Suridjan, Norbert Wild, Pieter J. Visser, Lars Bertram, Henrik Zetterberg, Kaj Blennow, Johan Gobom, Dan Western, Yun Ju Sung, Cynthia M. Carlsson, Sterling C. Johnson, Sanjay Asthana, Carlos Cruchaga, Betty M. Tijms, Corinne D. Engelman, Michael P. Snyder

## Abstract

The recently reported *TNIP1*/*GPX3* locus from AD GWAS studies was investigated. Using proteomics and other functional omics data, we identified evidence for a functional mechanism linking variants in this locus to decreased CSF GPX3 levels as AD progresses, suggesting a new potential target for intervention.

## Main

Genomic studies in Alzheimer’s disease (AD) have identified dozens of genetic associations^1–4^, but connecting variants with downstream drug targets is challenging. Individual functional validation to determine the consequences of mutations in implicated genes may be costly and time-consuming. In recent years, AD research studies have collected a variety of multiomics data on a large scale, including transcriptomics, proteomics, and metabolomics. These molecular data sets can provide key intermediate information linking genes to AD risk.

Several recent large-scale GWAS reported and discussed the rs871269 variant within an intron of the *TNIP1* gene (chromosome [chr] 5, base pair 151052827; all coordinates given in GRCh38 unless otherwise noted) as a protective variant for AD, with an odds ratio for AD case status of 0.96 (0.95-0.97) for the T allele, according to the Bellenguez et al. 2022 meta-analysis^5,6^. Wightman et al. additionally mention the rs34294852 variant (chr5:151053886), which is in low linkage disequilibrium (LD) with rs871269 in the East Asian (R^2^ = 0.29) and American (R^2^ = 0.22) populations but in lower LD (R^2^ = 0.14) among Europeans^7^. In terms of the functional connection of *TNIP1* with AD, Wightman et al. noted that there was little previous discussion regarding *TNIP1* and AD, but that of the three genes in the region of the variant (*TNIP1, GPX3*, and *SLC36A1*), *TNIP1* seemed to have the strongest connection through its regulation by Bcl3 in mice microglia and the corresponding BCL3 gene, which had been associated with CSF amyloid-beta levels in humans. Bellenguez et al. noted the connection of *TNIP1* with TNF-alpha, which was connected with the linear ubiquitin chain assembly complex (LUBAC), which itself is implicated in AD-relevant biology, including inflammation, microglia, and autophagy. Here, we use proteomics data from three different AD cohorts and annotation databases to investigate an alternative functional relationship of this locus to AD through the *GPX3* gene.

Using newly generated mass spectrometry cerebrospinal fluid (CSF) proteomics data from the University of Wisconsin (mean age 66.1; 59.9% female; detailed description previously reported^8^), we searched for evidence whether either TNIP1 or GPX3 levels were associated with AD. Specifically, we conducted ANOVA analyses that examined the relationship of these two proteins in CSF with CSF amyloid- and CSF tau-defined categories of AD (total n = 137; 56 A-T-, 39 A+T-, and 42 A+T+; see Methods). We used these amyloid (A) and tau (T) categories since they are the major biomarkers and central to research-based definitions of the disease, where amyloid tends to change first followed by tau in the current model of AD (note that A-T+ is generally excluded as presumably non-AD pathology)^9,10^. TNIP1 was not been detected in the CSF in the University of Wisconsin cohorts, but GPX3 (after correction for age and sex; see Methods) showed a statistically significant difference across the AD continuum from A-T-to A+T-to A+T+ (ANOVA P = 1.5 × 10^−5^) with a significant decrease between the A-T- and A+T+ categories (t-test P = 1.3 × 10^−5^) (Figure 1a). GPX3 was also significantly associated with 8 of the 9 tested individual markers of neurodegeneration and neuroinflammation (the Aβ42/Aβ40 ratio, phosphorylated tau [ptau], the ptau/Aβ42 ratio, alpha-synuclein, neurofilament light chain [NFL], neurogranin, YKL-40, and soluble TREM2 [sTREM2]), in each case negatively correlated with the biomarker except for Aβ42/Aβ40 (Figure 1b).

**Figure 1:**
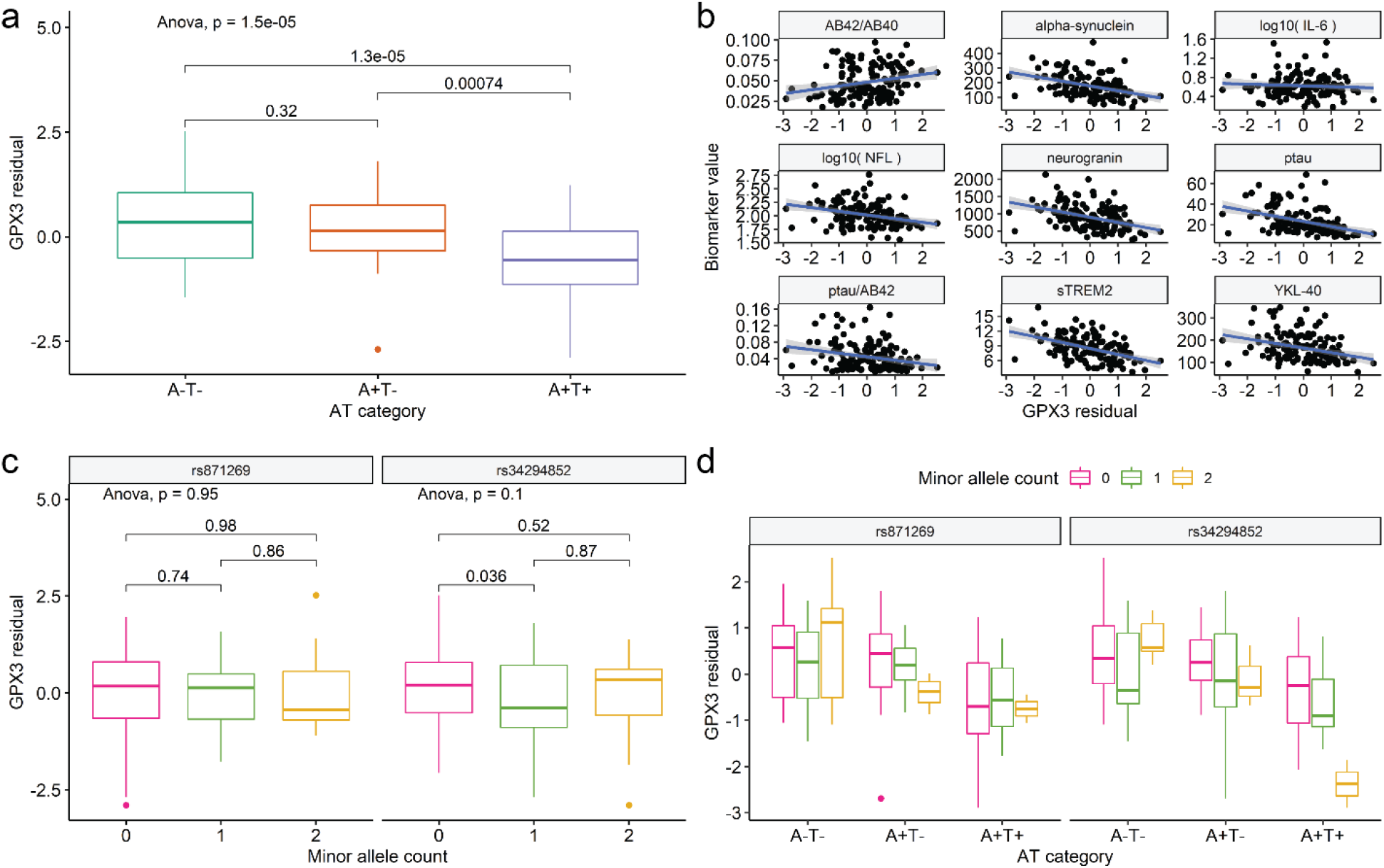
Associations of CSF GPX3 with AD-related measures in the Wisconsin ADRC and WRAP cohorts. **a)** The distribution of CSF GPX3 levels (after regressing out the effects of age and sex) significantly decreased across amyloid and tau (AT) positivity categories (n = 137). **b)** CSF GPX3 levels were significantly associated with all CSF biomarkers of neurodegeneration and neuroinflammation except for IL-6 (n = 137). In each case, GPX3 levels decreased as biomarker values indicated a worsening clinical profile. **c)** Across the cohorts (n = 137), no difference in GPX3 levels were observed by genotype of either AD-related variant. **d)** CSF GPX3 levels by both AT and genotype were summarized for both relevant SNPs at the *TNIP1*/*GPX3* locus. Among participants who were A+T+ (n = 42), CSF GPX3 levels were significantly decreased for homozygous recessive carriers of the rs34294852 allele (n = 5 among A-T-; n = 3 among A+T-; n = 2 among A+T+).

We then examined the genotype-mediated effects on the GPX3 trajectory. Visually and analytically (via ANOVA and pairwise t-tests), there were no differences in GPX3 levels by just the minor allele count of either SNP alone (ANOVA P values were 0.95 and 0.10 respectively across genotypes for rs871269 and rs34294852; Figure 1c), indicating that no differences in GPX3 level were present by genotype across the population. However, when we viewed these protein levels and genotypes along with AD biomarker categories, a dose-response relationship emerged for the rs34294852 genotype.

When GPX3 levels were analyzed as the outcome in a multiple linear regression with both AT category and SNP minor allele count (numeric coding) as predictors, a significant decrease in GPX3 levels per copy of the minor allele was observed (P = 0.041 for rs34294852; Figure 1d), suggesting that the *TNIP1*/*GPX3* locus-AD diagnosis relationship reported by GWAS might be mediated through changes in GPX3 level that might only become apparent when tau is abnormal in AD and particularly with homozygosity in the minor allele of rs34294852. In other words, changes in GPX3 levels might only be most evident for A+T+ individuals who are homozygous recessive for rs34294852.

Based on these results, we developed a hypothesis concerning the role of GPX3 in AD. GPX3 is a secreted glutathione peroxidase that works to protect the body from oxidative damage by reducing hydroperoxides^11^. AD has long been linked to oxidative stress, as have aging processes in general^12,13^. The observed decline in GPX3 levels in individuals with AD might reflect insufficient or deteriorating capacity to manage oxidative stress by those with AD. This effect might be exacerbated among carriers of the minor allele of rs34294852, which presumably decreases expression of GPX3 by affecting an enhancer region interacting with the *GPX3* gene. Given the rarity of the minor allele, the subtlety of the GWAS signal, and the presence of the SNP-protein association only in the context of AD, this mechanism might be hard to detect and perhaps only then under certain disease conditions (Figure 2).

**Figure 2:**
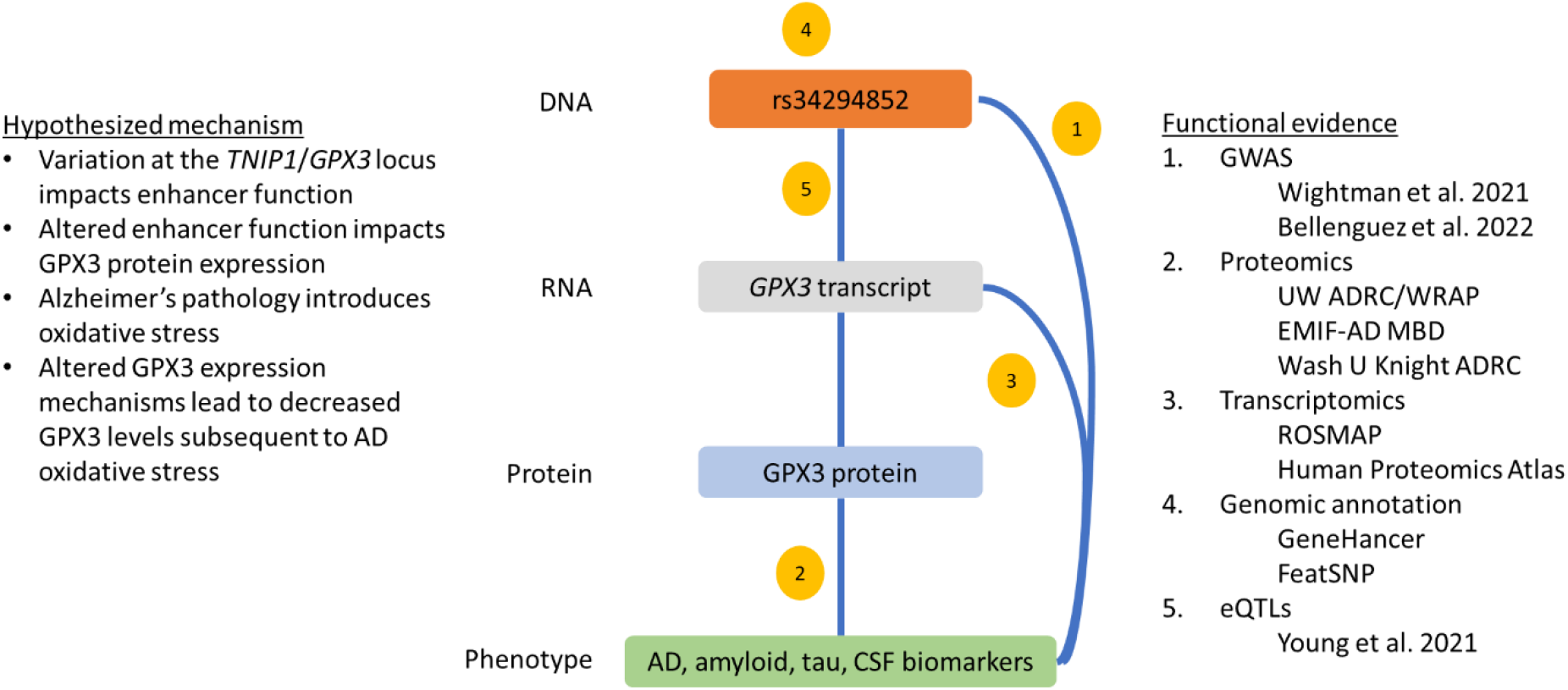
Proposed functional mechanism of the *TNIP1* locus in AD. Our hypothesis for a functional mechanism connecting the variant rs34294852 within *TNIP1* to AD outcomes is overlaid onto a map of the major omics data and the central dogma of biology. Major lines of post-GWAS functional evidence supporting this hypothesis are summarized in the right-hand list.

Were this hypothesis true, we would expect to see concordant evidence from AD transcriptomics experiments. First, we sought to understand which central nervous system cell types express *GPX3* in general. Across the cell types in the Human Protein Atlas, *GPX3* is most highly expressed in the proximal tubular cells of the kidney (20125.9 nTPM) and the Müller glia cells of the eye (5019.8 nTPM). Among brain tissues, the overall expression was lower, but microglia had the highest expression (3.7 nTPM) among the non-neuronal cell types, followed by astrocytes (1.0 nTPM) and oligodendrocytes (0.4 nTPM) (Supplementary Figure 1)^14^. Within an AD cohort, where GPX3 might be more relevant given the heterogeneity of the microglia transcriptome^15^, RNA-seq data from the prefrontal cortex from the Religious Orders Study Memory and Aging Project (ROSMAP) showed that *GPX3* transcript levels decreased in the prefrontal cortex from controls to AD diagnosis (P = 7 × 10^−6^; Supplementary Figure 2), but not in other brain regions^16^.

We also searched for evidence from functional genomics and transcriptomics experiments supporting a relationship between rs34294852 and *GPX3* transcription. In terms of genome functional annotation, rs34294852 is located within the 6^th^ intron of *TNIP1* and downstream of the enhancer region GH05J151051 from GeneHancer^17,18^. Variant rs34294852 is predicted to alter the binding of the transcription factor MZF1 according to FeatSNP^19^, which is a database that aggregates brain-specific epigenetic data to examine the effects of genetic variants. In terms of expression quantitative trait loci (eQTL) data, variant rs34294852 is an eQTL for TNIP1 and GPX3 in blood, and it is also an eQTL for TNIP1 in monocytes and neutrophils and for GPX3 for neutrophils^20–22^. Recognizing the relatively higher expression of GPX3 in microglia compared to other non-neuronal cell types and the implications of microglia in AD, we investigated recent eQTL data for microglia and found that rs34294852 was an eQTL for GPX3 (P = 0.038)^23^.

With the evidence from the original AD GWAS and our follow-up CSF proteomics and biomarkers experiments, these additional functional and molecular data supported our central hypothesis: genetic variation at the *TNIP1*/*GPX3* locus impacts expression of *GPX3* through impaired enhancer activity that in turn exacerbates the observed decrease in GPX3 levels as amyloid and ptau accumulate in AD progression. In cohort studies, this pathway might be difficult to detect for several reasons: 1) the original GWAS effect was small; 2) the pathway might only be relevant to a subset of cell types (e.g., microglia); and 3) and the effect might be harder to observe for homozygous dominant or heterozygous rs34294852 genotypes. Moreover, the effect allele for rs34294852 (C) has a minor allele frequency ranging from 0.16 for East Asians on the low end to 0.25 for African/African Americans on the high end^7^, which makes homozygous recessive individuals fairly uncommon, as was the case in the data sets analyzed here. Indeed, our attempts to replicate the proteomics signal with other population cohorts were unsuccessful. Using data from the European Medical Information Framework-Alzheimer’s Disease Multimodal Biomarker Discovery (EMIF-AD MBD) study (n = 242 participants), we performed an ANOVA analysis to see if GPX3 levels were different by AT category. No such difference was observed (P = 0.96), even when stratified by rs34294852 genotypes (Supplementary Figure 3) or controlling for age, sex, or study site. Similarly, no GPX3-AD association was observed using the Knight ADRC’s discovery and replication CSF proteomics data sets at Washington University in St. Louis (n = 1,168 and 597 participants, respectively). The differential abundance of GPX3 levels was not significantly different between A+T+ and A-T-individuals (P = 0.90 and P = 0.51, respectively) (Supplementary Figure 4). Differences in populations, proteomics technologies (mass spectrometry vs. aptamer-based), and amyloid and tau phenotyping might have affected the replication in addition to the other challenges noted above.

Nevertheless, the observed multiomic evidence connecting variation at this locus to AD through GPX3 expression, combined with our understanding of oxidative stress in AD and GPX proteins’ role in combating such stress, provide a compelling hypothesis for an alternative functional mechanism at this locus that may occur in concert with any effects through TNIP1. Several functional experiments would be a reasonable next step in exploring this hypothesis: 1) using a gene-editing experiment to validate the impact of genetic variation at rs34294852 on *GPX3* expression; 2) using proteomics analysis on a microglia cell line to validate GPX3 expression; and 3) using a cell or organismal model of AD to assess the longitudinal expression of GPX proteins in connection with oxidative stress burden and its relationship to changes in tau level. More broadly, the post-GWAS analyses here demonstrate the utility of multiomic cohort data in the investigation of GWAS loci and their mechanisms of action, which can lead to new insights into Alzheimer’s disease with the ultimate goal of identifying new therapeutic targets.

## Methods

### University of Wisconsin ADRC and WRAP cohorts

The discovery proteomics data set came from the University of Wisconsin Alzheimer’s Disease Research Center (ADRC)^24^ and Wisconsin Registry for Alzheimer’s Prevention (WRAP)^25^ cohorts, which have been described in detail previously^8^. This study was performed as part of the GeneRations Of WRAP (GROW) study, which was approved by the University of Wisconsin Health Sciences Institutional Review Board. Participants in the ADRC and WRAP studies provided written informed consent. Briefly, the data used here came from CSF samples taken from lumbar punctures (LPs) performed on middle- and older-aged participants in the ADRC and WRAP cohorts. All CSF samples were assayed between March 2019 and January 2020 at the Clinical Neurochemistry Laboratory at the University of Gothenburg. CSF biomarkers were assayed using the NeuroToolKit (NTK) (Roche Diagnostics International Ltd, Rotkreuz, Switzerland), a panel of automated Elecsys® and robust prototype immunoassays designed to generate reliable biomarker data that can be compared across cohorts.

Measurements with the following immunoassays were performed on a cobas e 601 analyzer (Roche Diagnostics International Ltd, Rotkreuz, Switzerland): Elecsys β-amyloid (1–42) CSF (Aβ42), Elecsys Phospho-Tau (181P) CSF (ptau), and Elecsys Total-Tau CSF, β-amyloid (1–40) CSF (Aβ40), and interleukin-6 (IL-6). The remaining NTK panel was assayed on a cobas e 411 analyzer (Roche Diagnostics International Ltd, Rotkreuz, Switzerland), including markers of synaptic damage and neuronal degeneration (neurogranin, neurofilament light protein [NFL], and alpha-synuclein) and markers of glial activation (chitinase-3-like protein 1 [YKL-40] and soluble triggering receptor expressed on myeloid cells 2 [sTREM2]). Participants were categorized as amyloid positive (A+) and tau positive (T+) based on the CSF amyloid (Aβ42/Aβ40 threshold = 0.046) and tau (ptau threshold = 24.8 pg/mL) measurements, as determined through previous work^26^. The fourth possible category of amyloid negative and tau positive (A-T+) was not included in this study as these samples were considered to represent non-AD pathological change^9^. A total of nine established CSF biomarkers for AD were analyzed in this study: the Aβ42/Aβ40 ratio, ptau (pg/mL), the ptau/Aβ42 ratio, NFL (pg/mL), alpha-synuclein (pg/mL), neurogranin (pg/mL), YKL-40 (ng/mL), sTREM2 (ng/mL), and IL-6 (pg/mL). NFL and IL-6 were log_10_-transformed to better normalize them due to right skew.

Genotyping was performed on these participants using DNA from whole blood samples. ADRC samples genotyped by the Alzheimer’s Disease Genetics Consortium (ADGC) at the National Alzheimer’s Coordinating Center (NACC) using the Illumina HumanOmniExpress-12v1_A, Infinium HumanOmniExpressExome-8 v1-2a, or Infinium Global Screening Array v1-0 (GSAMD-24v1-0_20011747_A1) BeadChip assay, while WRAP samples were genotyped with the Illumina Multi-Ethnic Genotyping Array at the University of Wisconsin Biotechnology Center^27^. Strict quality control (QC) steps and imputation to the Michigan Imputation Server^28^ and the Haplotype Reference Consortium (HRC) reference panel^29^ were performed. All individuals used in this study were of European ancestry. Throughout this paper, genomic coordinates are referred to by the GRCh38 genome build.

Proteomics data were generated on the CSF samples using an in-house single-shot nano-liquid chromatography-tandem mass spectrometry (nLC-MS/MS) method^8,30^ and the raw data quantified with MaxQuant^31,32^ using fast LFQ and a full human proteome with isoforms downloaded from UniProt (downloaded June 14, 2017). Protein abundance data were extracted from the LFQ intensity from the “proteinGroups.txt” output file. Strict QC steps were applied to the data, yielding a final set of 915 quantified proteins for the 137 participants. All time-variable data (age, proteomics, CSF biomarkers) were quantified on the same LP sample per participant; there was no sample time discrepancy between any of the data points for an individual.

The effects of age at sample LP and sex were first regressed out in a linear regression model (GPX3 ∼ age + sex), leaving the residuals for GPX3, which was the main variable of analysis in this data set. First, an ANOVA analysis was performed across the three AT-defined categories to look for a difference in GPX3 level, with pairwise t-tests also performed for each pair of AT categories and the results visualized with box plots. Second, GPX3 residual-biomarker associations were estimated for each biomarker separately using a linear regression model. A Bonferroni correction for the number of biomarkers tested (P = 0.05 / 9 = 0.0056) was used as the threshold for significance. Each biomarker was plotted against the GPX3 residuals, with a fitted regression line showing the relationship.

For genotype-stratified analysis, the box plots of GPX3 by AT category were repeated but with additional stratification by the count of minor alleles (T allele) for rs34294852. A linear regression was also performed with GPX3 residuals regressed on an indicator variable for A+T-, an indicator for A+T+, and the minor allele count for the variant (coded 0/1/2).

### GPX3 eQTL analysis

Microglia eQTL summary statistics were accessed through the European Genome-Phenome Archive (EGAD00001005736) and the Wellcome Sanger Institute Data Access Committee. All sequence datasets were aligned to human genome assembly GRCh38. Simple linear regression was used to map eQTLs with twenty-five principal components (PCs). The eQTL associations for *GPX3* were extracted for the analysis in this paper. Study designs and method details of microglia eQTL mapping have been described elsewhere^23^.

### EMIF-AD MBD cohort

The European Medical Information Framework for Alzheimer’s Disease Multimodal Biomarker Discovery (EMIF-AD MBD) study is a European multicenter study that aims to identify novel biomarkers for diagnosis and prognosis in the predementia stages of AD^33^. EMIF-AD MBD combines existing clinical data and samples of 1,218 individuals with normal cognition (controls), mild cognitive impairment (MCI), or mild dementia from prospective cohort studies. The study collected baseline clinical data, MRI scans, plasma, DNA, CSF samples, and follow-up diagnosis.

DNA was extracted locally at the collection site (n = 805) or from whole blood (n = 148) using QIAamp DNA Blood Mini Kit (QIAGEN GmbH, Hilden, Germany) at the University of Lübeck. Genetic samples were genotyped with the Illumina Global Screening array (GSA) custom content (Illumina, Incl) at the Institute of Clinical and Medical Biology (UKSH, Campus-Kiel). Quality control and imputation (HRC panel^29^) have been described in depth elsewhere^34,35^.

CSF was collected with lumbar puncture, as described previously^33^. CSF Aβ^1-42^ and total tau were measured locally with INNOTEST ELISA or INNOBIA AlzBio3 INNOBIA AlzBio3 (Fujirebio, Ghent, Belgium). Cut-offs for Aβ^1-42^ and total tau were cohort-specific in EMIF-AD. Because EMIF-AD centers had used different approaches to determine Aβ^1-42^ cut-offs, potentially leading to center-specific bias, we determined for each cohort specific cut-offs using unbiased Gaussian mixture modelling^36^. CSF GPX3 proteins levels were measured using tandem mass tag (TMT) mass spectrometry and have been deposited to the ProteomeXchange Consortium via the PRIDE partner repository with the dataset identifier 10.6019/PXD019910^37^. GPX3 CSF protein levels were Z-score normalized with controls with normal Aβ^1-42^ and normal total tau as reference group. A full description on the quality check and normalization procedures have been described elsewhere^34,38,39^.

A total of n = 242 subjects had genetic, AT category (CSF Aβ^1-42^ and total tau) and CSF proteomic data (i.e., GPX3) available (n = 242). Since normal CSF amyloid and abnormal CSF total tau (A-T+) probably reflects neurodegenerative disorders other than AD, we excluded subjects with A-T+ (n = 7) from further analysis, resulting in a total of n = 235 subjects (age in years mean (SD): 66.9 (8.1); female n (%): 127 (54%); AT category: 58 A-T-, 59 A+T-, 118 A+T+; diagnostic status: 105 controls, 64 MCI, 66 AD).

GWAS analysis on GPX3 CSF levels was performed using PLINK software (v1.9) with covariate adjustment for population structure (PC1-3), age, and sex^40^. ANOVA analysis was performed to examine CSF GPX3 differences across AT categories, including separate analyses that additionally included age and sex or age, sex, and study site as covariates.

Written informed consent was obtained from all participants or surrogates, and the procedures for this study were approved by the institutional review boards of all participating institutions, including the following (see Bos et al 2018 for full listing^33^): Aristotle University of Thessaloniki Medical School Ethics Committee; Ethics Committee of the Medical Faculty Mannheim, University of Heidelberg; Ethic and Clinical Research Committee Donostia; Ethics committee Inserm and Aix Marseille University; The Healthcare Ethics Committee of the Hospital Clínic; Central Clinical Research and Clinical Trials Unit (UICEC Sant Pau); INSERM Ethical Committee; Ethic Committee of the IRCCS San Giovanni di Dio FBF; Comitato Etico IRCCS Pascale - Napoli; Ethics Committee at Karolinska Institutet; Ethische commissie onderzoek UZ/KU Leuven; Research Ethics Committee Lausanne University Hospital; Medical ethical committee Maastricht University Medical Center; Committee on Health Research Ethics, Region of Denmark; Ethics committee of Mediterranean University; University of Lille Ethics committee; Ethical Committee at the Medical Faculty, Leipzig University; Ethical Committee at the Medical Faculty, University Hospital Essen; Ethics committee University of Antwerp; Ethical Committee of University of Genoa; Ethics Committee, University of Gothenburg; Human ethics Committee of the University of Perugia; and the Medical ethics committee VU Medical Center.

### Washington University in St. Louis cohorts

A total of 7,584 analytes were measured across 3,065 samples using SomaLogic’s Somascan platform. Participants were enrolled in the Memory and Aging Project (MAP) at the Knight Alzheimer’s Disease Research Center (Knight ADRC; n = 948), Alzheimer’s Disease Neuroimaging Initiative (ADNI; n = 758), the Dominantly Inherited Alzheimer Network (DIAN; n = 495), Pau (n = 232), and Ruiz (n = 632) studies. All participants provided informed consent to allow their data and biospecimens to be included. The study was approved by an Institutional Review Board at Washington University School of Medicine in St. Louis.

The expression level of 7,584 proteins was measured using a multiplexed, single-stranded DNA aptamer assay developed by SomaLogic. The protein levels were reported as relative units of intensity (RFU or Relative Fluorescence Unit). Initial data normalization was performed by SomaLogic using hybridization controls for intra-plate and median signal to account for inter-plate variances^41,42^ as well as normalization against an external reference to control for biological variances. A stringent quality check^43^ (QC) was performed in the normalized data matrix. At the end of the QC, 316 analytes and 83 subjects were removed, resulting in a final matrix with 7,268 analytes and 2,982 subjects.

CSF samples were collected in the morning after an overnight fast, processed, and stored at -80 °C. CSF data processing for each cohort are described in detail in the respective studies^44,45^. For CSF samples, case-control status was determined by the Clinical Dementia Rating (CDR) at the time of lumbar puncture. For this study, the CSF Aβ42 (A) and pTau181 (T) levels were used to perform the AT classification and biomarker positivity and negativity were used to perform the differential abundance analysis. These log-normalized CSF biomarker levels (AT) were used for dichotomizing each participant into biomarker positive (case) and negative (control), as previously described^44^.

A multiple linear regression model was used to identify differentially abundant analytes between discovery (MAP and Ruiz; 497 AT+ and 671 AT-participants) and replication cohorts (ADNI and Pau; 360 AT+ and 237 AT-participants) where age, gender, plate ID, and the first two surrogate variables (SVs) were used as the covariates.

## Data Availability

The data sets analyzed from the Wisconsin ADRC and WRAP studies may be requested at https://www.adrc.wisc.edu/apply-resources. Microglia eQTL summary statistics may be requested through the European Genome-Phenome Archive (EGAD00001005736) and the Wellcome Sanger Institute Data Access Committee. The EMIF-AD proteomics data may be requested from the ProteomeXchange Consortium via the PRIDE partner repository with the dataset identifier 10.6019/PXD019910. The Knight ADRC proteomic data is available at NIAGADS: NG00102 collection and can be interactively explored at http://ngi.pub:3838/ONTIME_Proteomics/.

http://ngi.pub:3838/ONTIME_Proteomics/

## Supplementary Figures

Supplementary Figure 1: *GPX3* transcript expression levels in different brain cell types

Supplementary Figure 2: *GPX3* transcript expression levels across different brain regions

Supplementary Figure 3: CSF GPX3 levels by AT category and rs34294852 genotype in the EMIF-AD MBD cohort

Supplementary Figure 4: CSF GPX3 levels by AT category in the MAP/Ruiz and ADNI/Pau cohorts

## Acknowledgments

We would like to thank the participants and staff of the WRAP, Wisconsin ADRC, EMIF-AD MBD, and Knight ADRC. Without their efforts this research would not be possible.

## Funding

This research is supported by National Institutes of Health (NIH) grants R01AG27161 (Wisconsin Registry for Alzheimer Prevention: Biomarkers of Preclinical AD), R01AG054047 (Genomic and Metabolomic Data Integration in a Longitudinal Cohort at Risk for Alzheimer’s Disease), P41GM108538 (National Center for Quantitative Biology of Complex Systems), R01AG037639 (White Matter Degeneration: Biomarkers in Preclinical Alzheimer’s Disease), R01AG021155 (The Longitudinal Course of Imaging Biomarkers in People at Risk of AD), R21AG067092 (Identifying Metabolomic Risk Factors in Plasma and CSF for Alzheimer’s Disease), and P50AG033514 and P30AG062715 (Wisconsin Alzheimer’s Disease Research Center Grant), the Clinical and Translational Science Award (CTSA) program through the NIH National Center for Advancing Translational Sciences (NCATS) grant UL1TR000427, and the University of Wisconsin-Madison Office of the Vice Chancellor for Research and Graduate Education with funding from the Wisconsin Alumni Research Foundation. Computational resources were supported by a core grant to the Center for Demography and Ecology at the University of Wisconsin-Madison (P2CHD047873). We also acknowledge use of the facilities of the Center for Demography of Health and Aging at the University of Wisconsin-Madison, funded by NIA Center grant P30AG017266. Author LMR was funded by the Memorabel fellowship “Identifying biological and clinical relevance of (epi)genetic risk factors in sporadic FTD” (ZonMW project number: 10510022110012). Author YKD was supported by a training grant from the National Institute on Aging (T32AG000213). Author PJV was supported by grants from the European Commission, IMI (AMYPAD: 115952; RADAR-AD: 806999; and EPND: 101034344) and the ZonMW (Redefining Alzheimer’s disease, #733050824736, and NCDC, #73305095005). Author HZ is a Wallenberg Scholar supported by grants from the Swedish Research Council (#2018-02532), the European Research Council (#101053962), Swedish State Support for Clinical Research (#ALFGBG-720931), the Alzheimer Drug Discovery Foundation (ADDF), USA (#201809-2016862), the AD Strategic Fund and the Alzheimer’s Association (#ADSF-21-831376-C, #ADSF-21-831381-C and #ADSF-21-831377-C), the Olav Thon Foundation, the Erling-Persson Family Foundation, Stiftelsen för Gamla Tjänarinnor, Hjärnfonden, Sweden (#FO2019-0228), the European Union’s Horizon 2020 research and innovation programme under the Marie Skłodowska-Curie grant agreement No 860197 (MIRIADE), and the UK Dementia Research Institute at UCL (#UKDRI-1003). Author KB is supported by the Swedish Research Council (#2017-00915), the Swedish Alzheimer Foundation (#AF-930351, #AF-939721, and #AF-968270), Hjärnfonden, Sweden (#FO2017-0243 and #ALZ2022-0006), the Swedish state under the agreement between the Swedish government and the County Councils, the ALF-agreement (#ALFGBG-715986 and #ALFGBG-965240), and the Alzheimer’s Association 2021 Zenith Award (ZEN-21-848495). The funders had no role in study design, data collection and analysis, decision to publish, or preparation of the manuscript. Author JG is supported by Alzheimerfonden (AF-930934) and the Foundation of Gamla Tjänarinnor. Author CC receives support from the National Institutes of Health (R01AG044546, R01AG064877, RF1AG053303, R01AG058501, U01AG058922, R01AG064614, 1RF1AG074007), and the Chuck Zuckerberg Initiative (CZI). The recruitment and clinical characterization of research participants at Washington University were supported by NIH P30AG066444, and P01AG003991. This work was supported by access to equipment made possible by the Hope Center for Neurological Disorders, the NeuroGenomics and Informatics Center (NGI: https://neurogenomics.wustl.edu/) and the Departments of Neurology and Psychiatry at Washington University School of Medicine. Authors LMR, PJV, and BMT are supported by the ZonMW Memorabel grant programma (#733050824), and author PJV is additionally supported by the Innovative Medicines Initiative Joint Undertaking under the EMIF grant agreement (#115372). ELECSYS, COBAS and COBAS E are trademarks of Roche. The Roche NeuroToolKit robust prototype assays are for investigational purposes only and are not approved for clinical use. We thank the University of Wisconsin Madison Biotechnology Center Gene Expression Center for providing Illumina Infinium genotyping services. The content is solely the responsibility of the authors and does not necessarily represent the official views of the NIH.

## Author contributions

Author DJP conceived the idea for the study; conducted the analyses of the various Wisconsin data sets; prepared the figures and tables; and led the writing of the manuscript. Author LMR conducted the analyses in the EMIF-AD MBD cohort. Author MPS helped conceive the idea for the study and interpret the results. Author MA conducted the analyses in the Knight ADRC cohort. Authors JL and QL provided the eQTL results in microglia. Author YKD contributed to the interpretation of the results. Authors HZ, KB, GK, IS, and NW contributed to the generation of the University of Wisconsin NTK CSF biomarker data set. Authors CDE, CMC, SCJ, SA, HZ, and KB contributed resources or funding. Authors CMC, SCJ, and SA assisted with the Wisconsin Alzheimer’s disease cohort studies and the data and analyses conducted there for this work. Authors DW, YS, and CC assisted with the Knight ADRC cohorts and the data and analyses conducted there for this work. Authors PJV, LB, HZ, KB, JG, and BMT assisted with the EMIF-AD MBD cohorts and the data and analyses conducted there for this work. All authors contributed to and critically reviewed the manuscript.

## Competing interests

Author CC receives research support from Biogen, EISAI, Alector, GSK and Parabon; these funders of the study had no role in the collection, analysis, or interpretation of data; in the writing of the report; or in the decision to submit the paper for publication. Author CC is a member of the advisory board of Vivid Genomics, Halia Therapeutics and ADx Healthcare. Author HZ has served at scientific advisory boards and/or as a consultant for Abbvie, Alector, Annexon, Apellis, Artery Therapeutics, AZTherapies, CogRx, Denali, Eisai, Nervgen, Novo Nordisk, Pinteon Therapeutics, Red Abbey Labs, Passage Bio, Roche, Samumed, Siemens Healthineers, Triplet Therapeutics, and Wave, has given lectures in symposia sponsored by Cellectricon, Fujirebio, Alzecure, Biogen, and Roche, and is a co-founder of Brain Biomarker Solutions in Gothenburg AB (BBS), which is a part of the GU Ventures Incubator Program. Author KB has served as a consultant, at advisory boards, or at data monitoring committees for Abcam, Axon, BioArctic, Biogen, Julius Clinical, Lilly, MagQu, Novartis, Roche Diagnostics, and Siemens Healthineers, and is a co-founder of Brain Biomarker Solutions in Gothenburg AB (BBS), which is a part of the GU Ventures Incubator Program. Author GK is a full-time employee of Roche Diagnostics GmbH. Author IS is a full-time employee and shareholder of Roche Diagnostics International Ltd. Author NW is a full-time employee of Roche Diagnostics GmbH. Author SCJ serves as a consultant to Roche Diagnostics and receives research funding from Cerveau Technologies. Author MPS is a cofounder and scientific advisor of Personalis, SensOmics, Qbio, January AI, Fodsel, Filtricine, Protos, RTHM, Iollo, Marble Therapeutics and Mirvie. He is a scientific advisor of Genapsys, Jupiter, Neuvivo, Swaza, and Mitrix. Other authors have no competing interests to declare.

## Notes

### Author Declarations

University of Wisconsin data sets: This study was performed as part of the GeneRations Of WRAP (GROW) study, which was approved by the University of Wisconsin Health Sciences Institutional Review Board. Participants in the ADRC and WRAP studies provided written informed consent. The institutional review boards of all participating institutions approved the procedures for this study. EMIF-AD MBD cohort data sets: Written informed consent was obtained from all participants or surrogates, and the procedures for this study were approved by the institutional review boards of all participating institutions, including the following (see Bos et al 2018 for full listing): Aristotle University of Thessaloniki Medical School Ethics Committee; Ethics Committee of the Medical Faculty Mannheim, University of Heidelberg; Ethic and Clinical Research Committee Donostia; Ethics committee Inserm and Aix Marseille University; The Healthcare Ethics Committee of the Hospital Clínic; Central Clinical Research and Clinical Trials Unit (UICEC Sant Pau); INSERM Ethical Committee; Ethic Committee of the IRCCS San Giovanni di Dio FBF; Comitato Etico IRCCS Pascale - Napoli; Ethics Committee at Karolinska Institutet; Ethische commissie onderzoek UZ/KU Leuven; Research Ethics Committee Lausanne University Hospital; Medical ethical committee Maastricht University Medical Center; Committee on Health Research Ethics, Region of Denmark; Ethics committee of Mediterranean University; University of Lille Ethics committee; Ethical Committee at the Medical Faculty, Leipzig University; Ethical Committee at the Medical Faculty, University Hospital Essen; Ethics committee University of Antwerp; Ethical Committee of University of Genoa; Ethics Committee, University of Gothenburg; Human ethics Committee of the University of Perugia; and the Medical ethics committee VU Medical Center. Washington University in St. Louis cohort data sets: The study was approved by an Institutional Review Board at Washington University School of Medicine in St. Louis.

