## Supplementary Figures for "Post-GWAS multiomic functional investigation of the *TNIP1* locus in Alzheimer’s disease implicates mediation through *GPX3*"

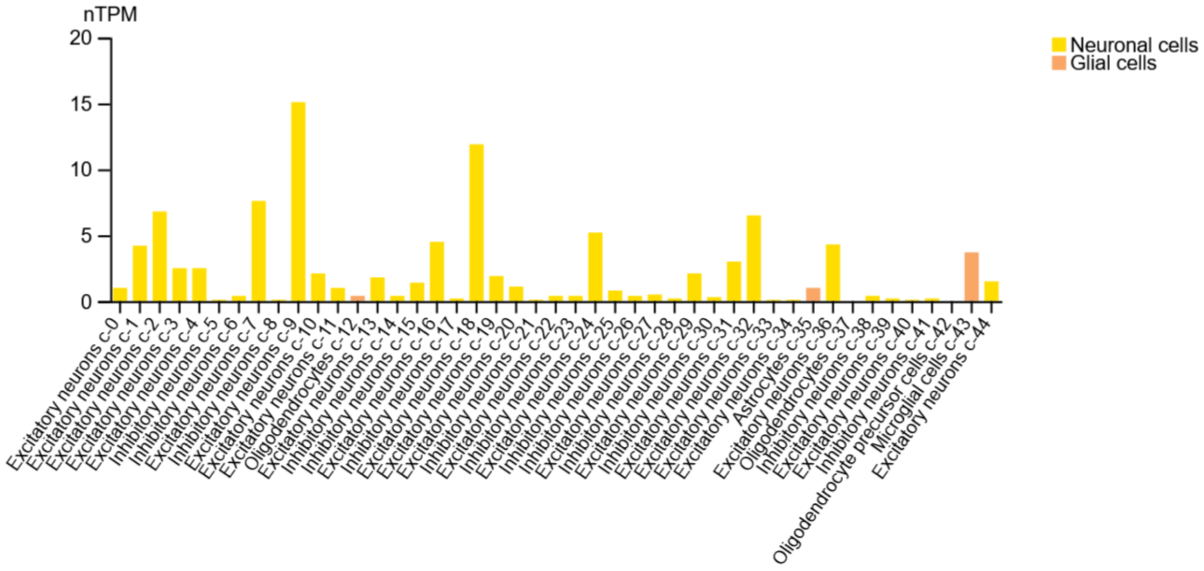

**Supplementary Figure 1.** ***GPX3*** **transcript expression levels in different brain cell types**

The transcript expression levels of *GPX3* in different brain cell types are shown. Data and the figure above were generated by the Human Protein Atlas and accessed on 4/12/2022 (https://v21.proteinatlas.org/ENSG00000211445-GPX3/single+cell+type) (see main manuscript for full citation). Among the non-neuronal cell types, microglia had the highest expression of *GPX3* (3.7 nTPM). nTPM stands for normalized transcripts per million.

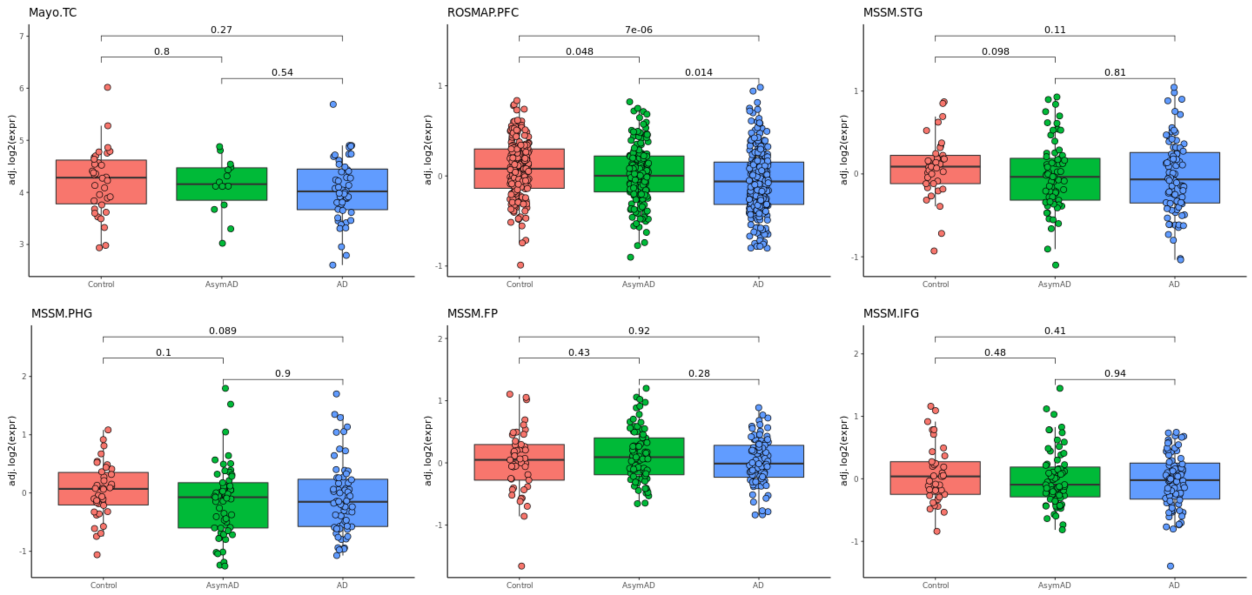

**Supplementary Figure 2. *GPX3* transcript expression levels across different brain regions**

The levels of *GPX3* transcripts measured in different brain regions across diagnoses of AD (control, asymptomatic AD [“AsymAD”], and AD) as reported by Morabito et al. 2020 in *Human Molecular Genetics* (accessed via http://swaruplab.bio.uci.edu:3838/bulkRNA/ on 4/15/2022). The following regions were analyzed: temporal cortex (TC), prefrontal cortex (PFC), superior temporal gyrus (STG), para-hippocampal gyrus (PHG), frontal pole (FP), and inferior frontal gyrus (IFG). Underlying data sets included the Mayo Clinic Brain Bank (Mayo), the Religious Orders Study and Memory and Aging Project (ROSMAP), and the Mount Sinai School of Medicine (MSSM).

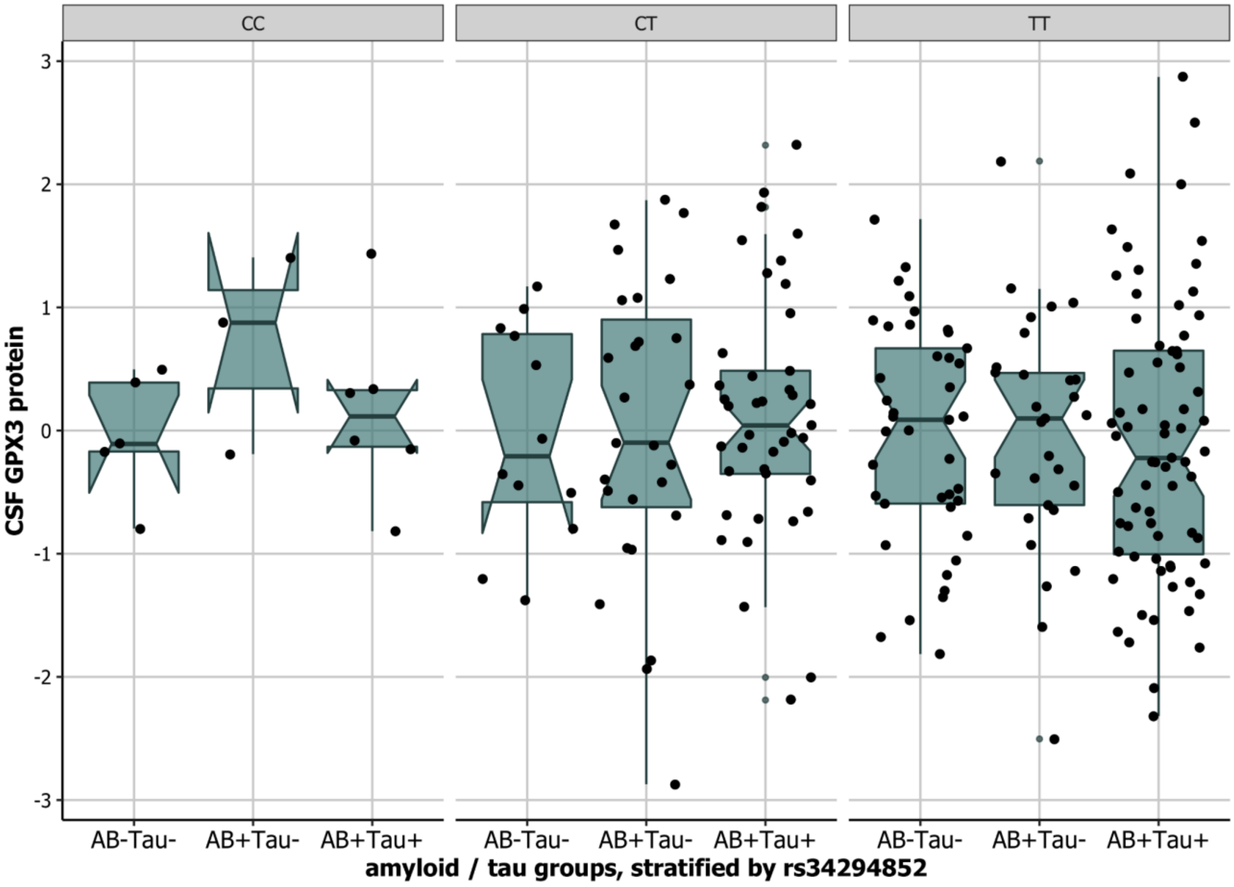

**Supplementary Figure 3. CSF GPX3 levels by AT category and rs34294852 genotype in the EMIF-AD MBD cohort**

The distribution of CSF GPX3 levels across AT-defined categories and by rs34294852 genotype is shown as measured in the EMIF-AD MBD cohort (n = 242). No statistically significant difference in GPX3 levels was observed from an ANOVA test ($P = 0.96$) nor any clear decline visually.

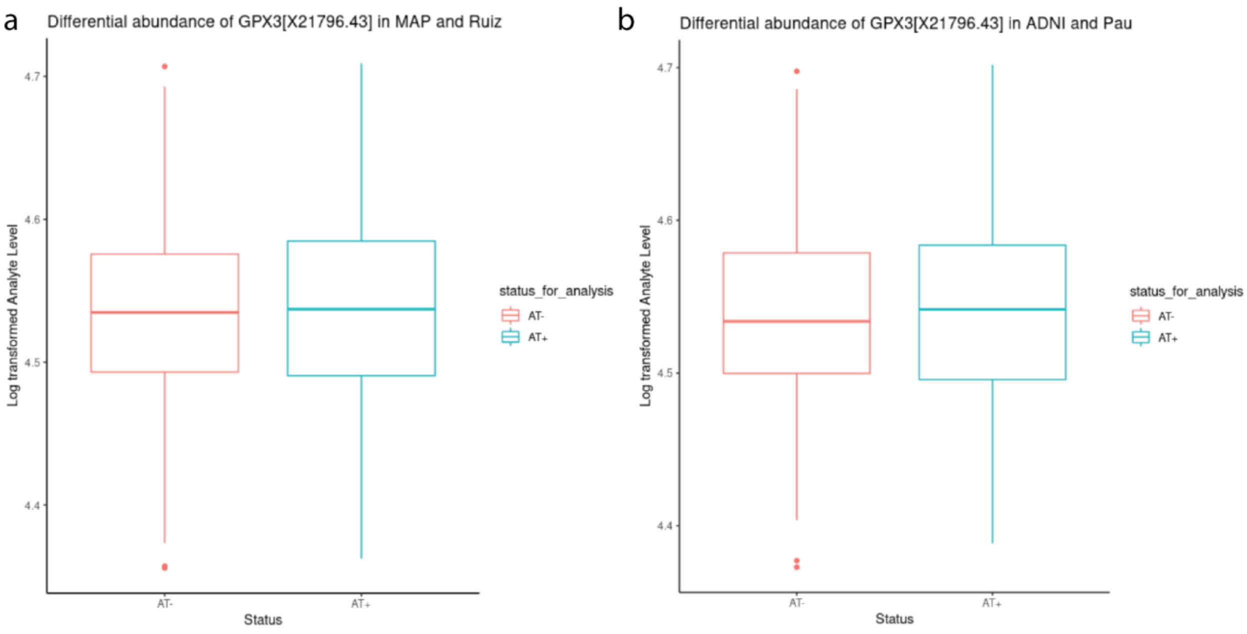

**Supplementary Figure 4. CSF GPX3 levels by AT category in the MAP/Ruiz and ADNI/Pau cohorts**

The distribution of CSF GPX3 levels between A-T- and A+T+ categories for two data sets analyzed by Washington University in St. Louis is shown. **a)** The discovery analysis data set (n = 1,168) distribution. **b)** The replication analysis data set distribution (n = 597). Neither data set showed a statistically significant difference in GPX3 values (P = 0.90 and P = 0.51, respectively).
